# Covid-19 trajectories – Monitoring pandemic in the worldwide context

**DOI:** 10.1101/2020.06.04.20120725

**Authors:** Henry Loeffler-Wirth, Maria Schmidt, Hans Binder

**Affiliations:** IZBI, Interdisciplinary Centre for Bioinformatics, Universität Leipzig, Härtelstr. 16 – 18, 04107 Leipzig, Germany

## Abstract

**Background:** Covid-19 pandemic is developing worldwide with common dynamics but also with partly marked differences between regions and countries. They are not completely understood, but presumably, provide one clue to find ways to mitigate epidemics until exit strategies to its eradication become available.

**Method:** We provide a monitoring tool available at www.izbi.de. It enables inspection of the dynamic state of the epidemic in 187 countries using trajectories. They visualize transmission and removal rates of the epidemic and this way bridge epi-curve tracking with modelling approaches.

**Results:** Examples were provided which characterize state of epidemic in different regions of the world in terms of fast and slow growing and decaying regimes and estimate associated rate factors. Basic spread of the disease associates with transmission between two individuals every two-three days on the average. Non-pharmaceutical interventions decrease this value to up to ten days where ‘complete lock down’ measures are required to stop the epidemic. Comparison of trajectories revealed marked differences between the countries regarding efficiency of measures taken against the epidemic. Trajectories also reveal marked country-specific dynamics of recovery and death rates.

**Conclusions:** The results presented refer to the pandemic state in May 2020 and can serve as ‘working instruction’ for timely monitoring using the interactive monitoring tool as a sort of ‘seismometer’ for the evaluation of the state of epidemic, e.g., the possible effect of measures taken in both, lock-down and lock-up directions. Comparison of trajectories between countries and regions will support developing hypotheses and models to better understand regional differences of dynamics of Covid-19.

## Introduction

Coronavirus disease (COVID-19) arrived in 187 countries with 5.5 Mio infections and more than 300,000 deaths worldwide so far (25^th^ May 2020). The disease affects almost all spheres of life, especially public health, economics and well-being. Present situation and near future lasting from months to one-two years (in worst-case more, in best-case less) will require coexistence with the virus until effective pharmaceutical countermeasures (medication, vaccine) are available and applicable [1]. This coexistence requires adjustment of a balance between a controllable low level of infections and maximum-possible levels of public life and economics.

Controlling the infection requires feedback loops sensitive to early and robust indications of secondary outbreak waves. This includes permanent surveillance of epidemiologic and medical indicators by testing programs, monitoring of case numbers and symptoms and forecasting methods on one hand, and suited ‘no-pharmacological intervention’ (NPIs) strategies on the other hand, to held the case numbers low (ideally further decreasing) to prevent secondary outbreaks.

Various ‘number-tracker’ tools are active (e.g., [2-5]). They mostly plot case numbers (infected, recovered, death) over time, usually on a country-by-country (or region-by-region) basis. As an illustration, we show the number of current infections and of Covid-19 related deaths as a function of time in selected countries (Figure 1). These ‘epi-curves’ reveal how the epidemic was expanding in time and space from China (end of 2019 and January 2020) via other Asian countries (South Korea, Iran) and Western Europe towards Eastern Europe, Amerika, and other parts of the world (February to April 2020). They also reveal different phases of epidemic, namely, an initial ‘take-off stage’, an ‘exponential growing stage’ followed by ‘slowed growth’, ‘turning into a decline’ and ‘decline’ [5]. Charting the outbreak day by day in each country and comparing them, e.g. by setting an arbitrary starting threshold of, for example, 100 infections, illustrates the succession of events as a global story [2]. For a straightforward evaluation simple measures are typically used such as doubling time of cases, reproduction numbers (mean number of people infected by a typical case) or the number of new infections per 100,000 citizens in a certain region, which all provide a limited snapshot-view with pro’s and con’s in different states of epidemic.

**Figure 1:**
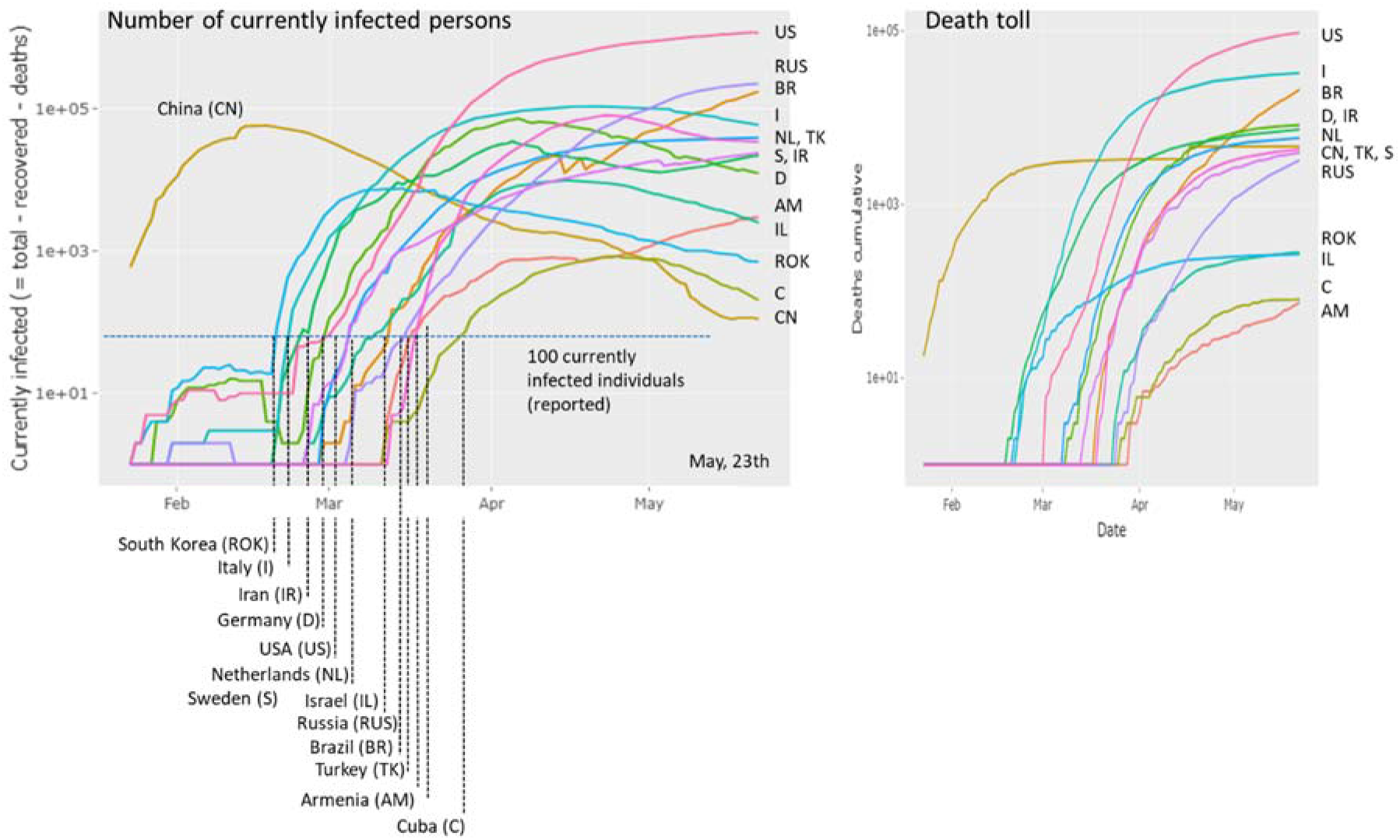
Covid-19 cases (left plot: currently infected, right plot: died individuals) in different countries as a function of date. The ‘100-cases per country’ threshold is crossed between end of February and end of March for the countries shown (except China). The time courses reflect growing (e.g., US, RUS, NL), slightly decaying (e.g., I, D, TK), strongly decaying (e.g., CH, ROK) regimes of epidemic or indications of bi-or multiphasic growth (e.g. AM, IR). The courses of the dead toll as a function of time reflect country-specific percentages of Covid-19 victims. The plots were generated in the Corona-viewer on a daily actualization-basis as described in the text.

As a next, more elaborated level, standard epidemic models provide a theoretically well-founded description of dynamics of disease incidence in terms of rate constants for transmission and recovery of Covid-19 and detailed infection-transmission ‘serial interval’ functions. Different models, mostly assuming a series of diseases states such as the ‘Susceptible-Infected-Removed’ (SIR) types (see below) have been used to describe ‘epi-curves’ of selected countries and regions under consideration of i) spatial heterogeneous outbreak and transmission scenarios, and ii) the effect of NPIs [6-21]. In case of the latter, models have been applied not only in retro-perspective but also to forecast epidemic in dependence on measures taken. Because of still limited knowledge about disease mechanisms and detailed data about its spread in the population forecasting either provides short-term extrapolations or hypothetical predictions of possible future scenarios as the result of different model assumptions.

We here provide the Covid-19 viewer, a monitoring tool which aims at bridging the temporal ‘epi-curve’ and the modelling levels. Our monitoring substitute the time-coordinate used in the epi-curves by infected cases (cumulative or current ones). The obtained trajectories then enable to visually estimate the dynamic state of epidemic in terms of simple shape characteristics such as slope, parallel shifts or turning points with direct relations to transmission and removal rates of the disease. Comparative analysis between trajectories of different countries enables to judge different scenarios of NPIs, population size, and social factors. Daily actualized data and interactive web-functionalities enable monitoring pandemic based on newest data. Our trajectory-approach is complemented by a series of simple model calculation which visualize the obtained trajectories for comparison with real ones.

The paper is organized as follows: In the results section we introduce and illustrate the different trajectories and plots available in the monitoring tool by showing examples from different countries of the world, which are thought to serve as worked examples referring to the actual state of pandemic in the second half of May 2020. The majority of plots shown in the publication were directly taken from the web-tool. The interested reader thus can actualize the data and/or chose countries of interest for similar views. We address the effect of NIPs in Europe, the spread of epidemic in Germany and compare mortalities between selected countries. The Materials and Methods section shortly explains the major functionalities. Details of the methods, model simulations and fits as well as supplementary figures were provided in the Supplement (Appendix).

## Methods and Data: Monitoring Covid-19 numbers

### Tool and data

The trajectory-monitoring tool (‘Covid-19 viewer’) was programmed as web application using the R-package ‘shiny’ [22]. It processes the number of newly infected and of removed (sum of recovered and died) individuals from 187 countries (and of Diamond Princess cruise liner with 712 cases) as provided by the Corona virus resource center of Johns Hopkins Medical University (‘World data’: https://systems.jhu.edu/research/public-health/ncov/) and from Robert-Koch-Institut (‘German-country’ data: https://www.rki.de/DE/Content/InfAZ/N7Neuartiges_Coronavirus/Fallzahlen.html). Data are daily updated.

### Availability

The tool is available via the websites of IZBI (www.izbi.de) and the Leipzig Health Atlas (https://www.health-atlas.de/models/28).

### Functionalities

The ‘Covid-19 viewer’ is an interactive tool to monitor the development of the pandemic in 187 countries and in the 16 German states using simple and intuitive plots (Figure 2, Appendix I). The tool is interactive and enables the user to select different presentations of data. The so-called ‘rise-fall’ trajectory was chosen as ‘standard visualization’. It shows the newly confirmed Covid-19 cases per country and per day (averaged over the past 7-days) as a function of accumulated total cases per country in double-logarithmic scale. The ‘rise-fall’ trajectory typically divides into a ‘rising’ exponential growth part reflecting growth of epidemic and a ‘falling’ decay regime due to counter measures and/or progressive immunization in the population. It allows estimating transmission and removal rates and reproduction numbers (Appendix I). The time range can be chosen and, as an illustration, pressing the ‘start animation’ button generates a movie of the dynamics of epidemic in the selected countries in terms of progressing rise-fall trajectories. The user can chose ‘custom’ trajectories to combine different numbers (infected or removed cases, deaths, daily or cumulative counts, Figure 1) along the coordinate axes for alternative views (use the hoover window for curve assignment and details such as date, numbers). Trajectories can be generated for all countries, groups of countries (use left-handed table for selection) or single countries. German states can be selected by choosing ‘Germany-state codes’. In addition to the standard plot, conventional time series plots show the different numbers (infected, removed, recovered, died as cumulative or per-day) as a function of date. The viewer offers standard browsing functionalities (zooming in and out, image download).

**Figure 2:**
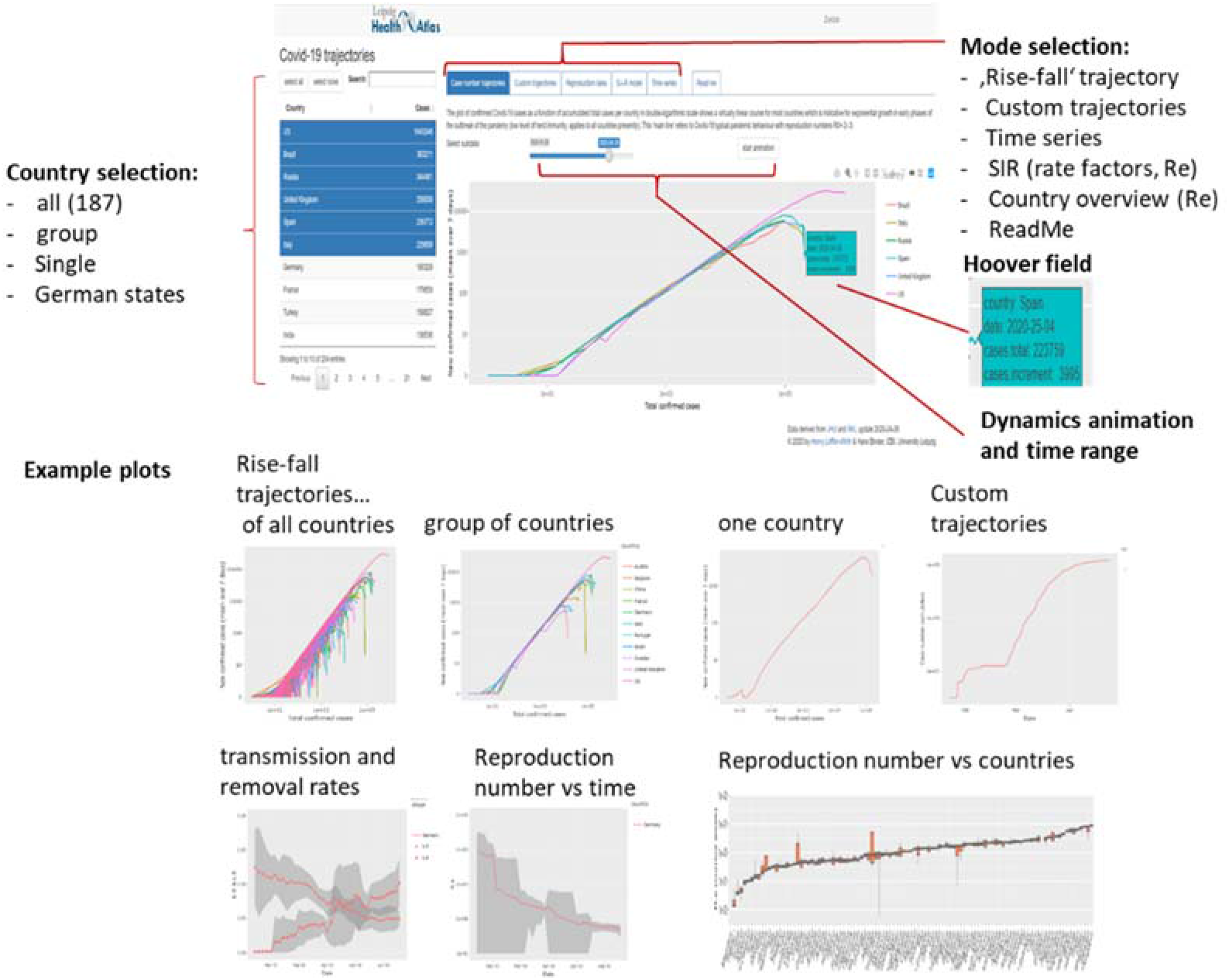
Covid-19 viewer: Screenshot with major functionalities indicated (above) and example plots (below).

### SIR model: Parameter estimation and visualization

The SIR (Susceptible-Infected – Removed) model provides a simple, adequate and straightforward interpretation of the data (see Figure 3 for illustration and Appendix II). It describes the disease as a sequence of three states, S (susceptible), I (infected) and R (removed), where infection proceeds via interactions between S and I individuals. Recovered individuals are assumed to get immunized. The respective numbers were reported by census systems, which can differ between countries, e.g. by counting only hospitalized individuals, counting died Covid-19 positive cases as not Covid-19 caused and/or referring to different test-frequencies. All case numbers must therefore be understood as ‘visible’, i.e. reported ones.

**Figure 3:**
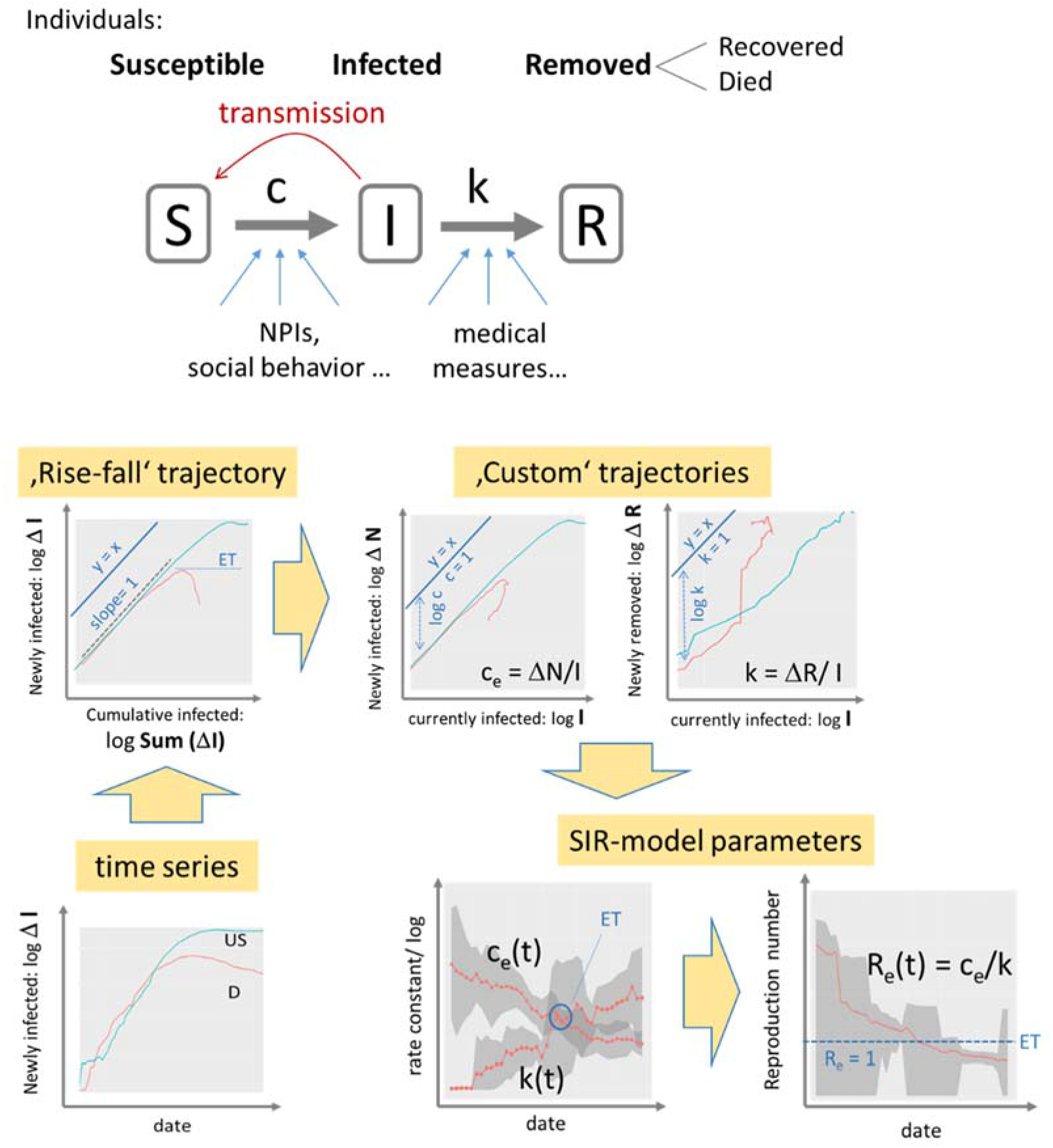
SIR model analysis: The SIR model assumes a sequence of three disease states (S, I, R, S+I+R=N, N population size) and transitions between them. Transmission and removal rates (numbers per day) are quantified in terms of the rate constants c and k, respectively. The time series of infected cases transforms into trajectories, which show daily changes of infected and removed cases as a function of cumulated or current infections in double logarithmic scale. Exponential growth is indicated by slope of unity while the transmission and removal rate factors refer to the vertical distance between the respective trajectory and the diagonal line (y= x). A downwards curved trajectory indicates decreased growth or eradication of epidemic (slope<0) with the epidemic threshold (ET: slope= 0) indicating stop of epidemic. Values of the factors c_e_ and k and of the reproduction number R_e_ are plotted as a function of date as running averages over a sliding window of seven days (the running standard deviation is shown as grey uncertainty strip). Example data are shown for Covid-19 case numbers in Germany (D, red curves) and USA (US, blue) from 27.04.2020. See text and Appendix I and II for details.

The rise-fall trajectories enables classification of the type of the growth and identification of the epidemic threshold (no growth). Custom trajectories allow to estimate time-dependent SIR model parameters such as the effective **transmission and removal rate factors**, c_e_(t) and k(t), respectively. Time courses of the rate factors were extracted from the local slopes of the trajectories (Appendix I and II). The ratio of the rate factors estimates the **effective reproduction number** R_e_(t) defined as the number of individuals who get contaminated by one infected person on the average. The time-dependent rate factors depend, in addition to the intrinsic properties of Covid-19 on a series of external factors such as Public Health Measures (<u>N</u>on-<u>P</u>harmaceutic <u>I</u>nterventions, NPIs) to slow down transmission of epidemics (affecting c_e_) and effective medical services after infection (affecting k).

In addition to the estimation of SIR parameters as described above, we performed least-squared fits of the trajectories where the daily numbers of newly infected and removed cases were calculated as a function of the cumulative number as predicted by the SIR model (Appendix II). The fits provide estimates of N_max_, the maximum cumulative number of infected cases, and of the rate constants.

## Results

### Monitoring the state of epidemic using ‘rise-fall’ trajectories

The ‘rise-fall’ trajectory plots the newly confirmed Covid-19 cases (averaged over a running 7 day windows) as a function of accumulated total cases per country in double-logarithmic scale. The ‘select all’ function shows the trajectories of all countries considered (Figure 4a). Overall, these double – logarithmic trajectories reveal two basic features: An initially linear increase with a slope of unity indicates exponential growth of epidemic. This linear regime is followed for many countries by a downwards turn which indicates slowing down of growth owing to NPIs ‘locking down’ infections and/or possibly also to progressing immunization of the population in later phases of epidemic leading to the depletion of the reservoir of susceptible individuals and/or other factors.

**Figure 4:**
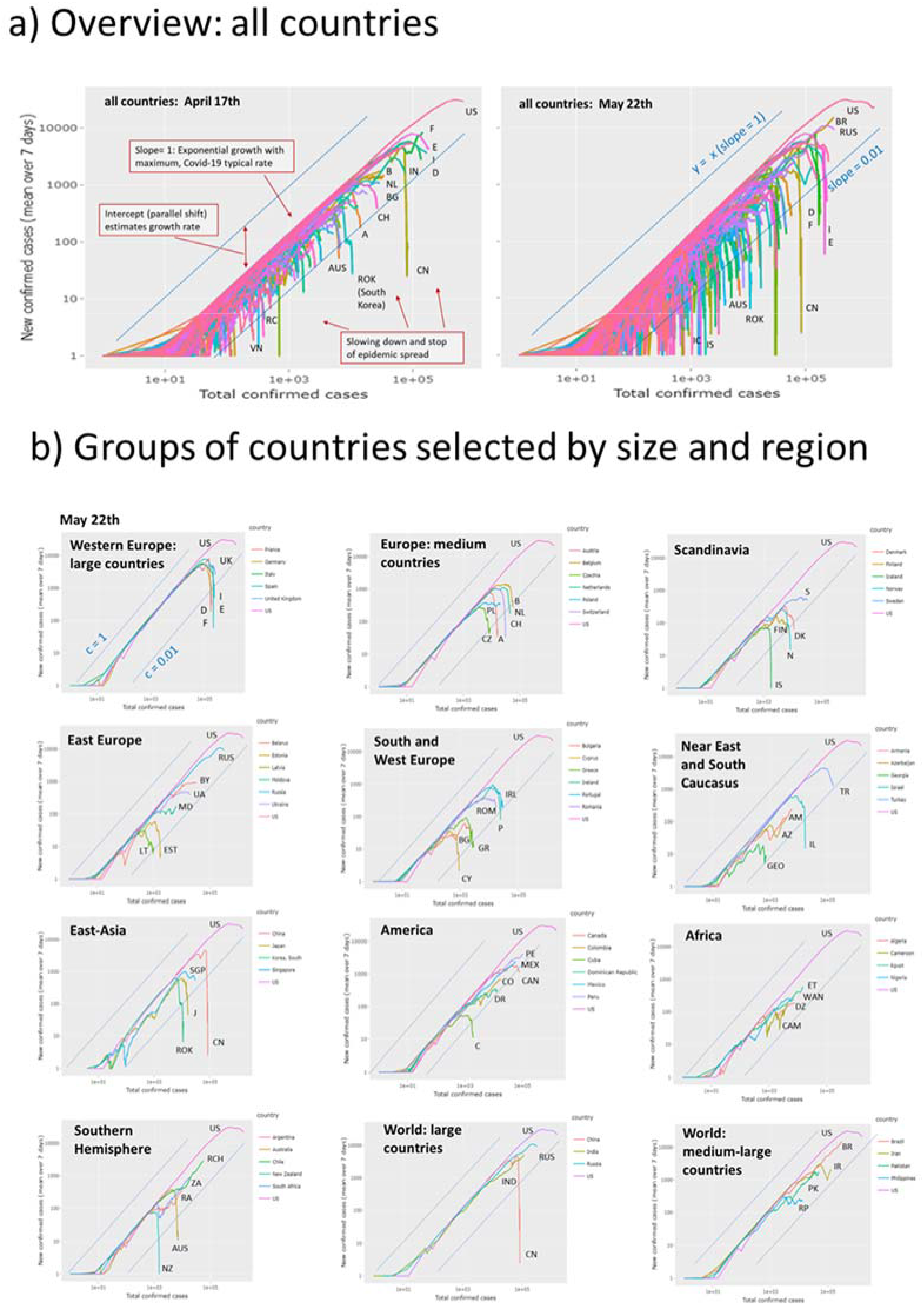
‘Rise-and-fall’ trajectories of Covid-19 pandemic: a) Trajectories of all 187 countries considered by Johns Hopkins Data Center plot new confirmed cases as a function of total cases in double logarithmic scale. They visualize the state of the epidemic with country-wise resolution. Exponential growth is reflected by a line parallel to the diagonal y= x (slope =1) followed by a fall reflecting slowing down of the epidemic. The slope equals the difference between the transmission and removal rate factors, c-k. The slope= 0.01 line consequently refers to the more as tenfold reduction of the maximum transmission rate observed for many countries in their linear rising part. With time (compare left and right plots) the number of countries crossing this line increases. b) Trajectories for groups of countries characterize the epidemic in countries of different population sizes and regions of the world. Trajectory of USA (with highest infection numbers worldwide) is shown for comparison in all plots. Letter codes for countries were chosen according to the international vehicle registration code (https://en.wikipedia.org/wiki/International_vehicle_registration_code).

The ‘rise-fall’ plots use the cumulative number of cases N as a robust measure of progressing epidemic in a population. Naturally, it is larger for countries with larger population sizes providing a larger overall reservoir for Covid-19 infections compared with smaller countries. Shape of the ‘rise-fall’ trajectories are however virtually independent of country size. The trajectories thus reflect intrinsic properties of epidemic in terms of its transmission and removal potential. The two sets of trajectories shown in the left and right part of Figure 4a refer to situation at April 17th 2020 and about six weeks later, respectively. For most countries, among them France, Italy, Spain and Germany, the trajectories turn into falling courses during this time and/or the falling parts further drop and intersects the ‘0.01 -slope’ line referring to a more than tenfold reduction of the transmission rate of epidemic (see below). These trends thus indicate decay of pandemic after the NPIs taken in most of countries. On the other hand, Brazil and Russia emerged to the countries with most cumulative cases after USA, with still growing case numbers.

Detailed inspection of the trajectories of sets of countries grouped roughly by size and geographic region reveals further details (Figure 4b, compare with Figure S 4 in Appendix III showing the respective trajectories recorded at April 29th). Most Western European countries of larger and medium size reached the decaying part in the first week of April 2020 (except Sweden and Great Britain) roughly two-three weeks after NPIs were taken in these countries. Countries from different parts of the world such as Austria, Iceland, South Korea, Australia, New Zealand and China reached low levels of new infections as indicated by strong vertical decays. Larger countries (e.g. Russia, India, Brazil, Pakistan) were in the rising part. Some countries show a two-phasic growth as indicated by the parallel right shift of linear regions in their growing part (e.g. Sweden, Denmark, Iran, Ukraine, Armenia) indicating that fast exponential growths are followed by slower phases due to reduced transmission rates (see below). Singapore and Japan show relatively slow growing phases with reduced rates and late turns into falling regimes while South Korea’s turn is very sharp presumably because of the ‘crash down’ measures taken there.

According to different regions and country sizes one finds that larger European countries (D, F, E, I, we use vehicle country codes for abbreviations of countries) except GB, selected middle-sized European countries (B, NL, CZ, CH, A) except PL and Scandinavian countries (DK, N, FIN, ISL) except S show a strong drop of epidemic. The exceptions are characterized by slowed-down growth (S, PL, GB). East Europe (RUS, UA, BY, MD), South Caucasus (AM, GEO, AZ), selected American (US, PE, MEX, CO, DR) and African (ET, WAN, CAM, DZ) countries all did not reach the turning point into the falling regime. Exception from these regions are CAN, C, the Baltic countries (EST, LAT), TK and IL. States from the Southern Hemisphere split into ones with stopped Epidemic (NZ, AUS) and ones with still spreading epidemic (RA, ZA, RCH), also seen for large (IND) and medium-sized countries (BR, PK, IR). Overall inspection of trajectories indicates that spreading pandemic shifted from East Asia and Middle and West Europe towards South America, South Asia and Africa. South Caucasus, Middle Asia (IR) and Eastern Europe show a heterogeneous picture.

Hence, the ‘rise-fall’ trajectories illustrate the current state of the epidemic and its developmental course with country-wise resolution. They enable monitoring the state in terms of differences and similarities between the countries and geographic regions revealing specifics and commons of epidemic spread: (i) A unique linear slope of most of the trajectories in the intermediate abscissa range is indicative for exponential growth in early phases of the outbreak of the pandemic (low level of immunity in the population). The nearly identical position of these lines refers to Covid-19 typical pandemic spread rate and maximum basic reproduction numbers R_0_ (Appendix II). (ii) Parallel, down-shifted lines suggest still exponential growth, however with reduced rates reflecting reduced effective reproduction numbers 1 < R_e_ < R_0_. In these countries (e.g., Sweden, Iran), the epidemic is not stopped. (iii) The ‘flattening’ of slope and downwards curvature seen, e.g. for most European countries such as Italy, Spain or Germany reflects slowing down growth owing to efficiency of NPIs and, possibly to a minor degree of progressive and significant immunization in the population. (iv) The sharp, virtually vertical drop of trajectories reflects the stop of epidemic observed, e.g. for China and South Korea, and after May 1st for New Zealand, Australia, and also part of European countries. (v) The different qualitative features of the trajectories are virtually independent of (population) size of the countries. ‘Smaller’ countries like Island, Cyprus, Armenia or Georgia show overall similar features such as linear rise, parallel shifts (Armenia), a maximum and steep falling parts (e.g., Island).

### Monitoring Covid-19 by custom trajectories, epi-curves and rate factors

The ‘rise-fall’ trajectory uses cumulative cases N along the abscissa as a robust measure of the extent of the epidemic. This number doesn’t consider the degree of recovery and thus it doesn’t reflect the current amount of infected cases (I). Custom trajectories make use of the independent number of removed cases (R) reported and plot cumulative, current and differential (per day) numbers in different combinations (Appendix II). Using the current number of infected cases (I= N – R) as x-axis one sees whether the extent of infection increases or it decays. While the rise-fall trajectory, ΔN-vs-N, tends asymptotically towards a maximum cumulative number of infections (N_max_) for each country, which reached the falling regime, the ΔN-vs-I trajectory turns from a growing I into a decaying branch at I_max_, the maximum number of infected individuals. These trajectories turn in clock-wise directions for most countries meaning that the rate factor of transmission of epidemic, c_e_(t), strongly decays (Appendix I and II). For example, Austria and Japan show full turns while the turns of Sweden and USA remain incomplete leading to less pronounced decays of the respective c_e_(t)-courses (Figure 5a, b). In contrast, the ΔR-vs-I trajectories turn typically in counter-clockwise direction referring to an increase of the removal rate factor as explicitly seen in the respective k(t) plots. The ratio of the effective transmission and of the removal rates then estimates the effective reproduction number as a function of time, R_e_(t) (Figure 5b). The trajectories of the countries selected for illustration reflect different types of trends such as strong and straight repression and stop of epidemic via reduction of transmission in Austria, reduced growth but still expanding epidemic in USA and Sweden or indications of a second wave of expanding epidemic in Iran. Here, the respective trajectories and plots of rate factors and of R_e_(t) show different aspects of the dynamic of the epidemic. For example, S and J are characterized by relatively low levels of rate factors compared with A and IR, a difference seen also in the parallel shifts of the respective trajectories. In Appendix II we find analogous differences between Western and South European countries (E, F, I, Figure S 3) compared with Middle European ones (D, A, CH), which suggest differences in the spread mechanism of Covid-19 and, possibly, also in the recovery dynamics. The removal rate obtained depends on the time-delay between infection and recovery, which is neglected our simple trajectory-approach (see also epicurves in Figure 5a). ‘Cumulative balance’ and ‘current case’ custom trajectories complete the visualization options: Lower levels of transmission and removal rates associate with such trajectories running closer to the diagonal. Overall, the trajectories enable tracing an epidemic in terms of case numbers reported directly be the census agencies of the respective countries. Derived numbers such as the rate factors and reproduction numbers ‘translate’ these numbers into features more directly describing the dynamics of the epidemic.

**Figure 5:**
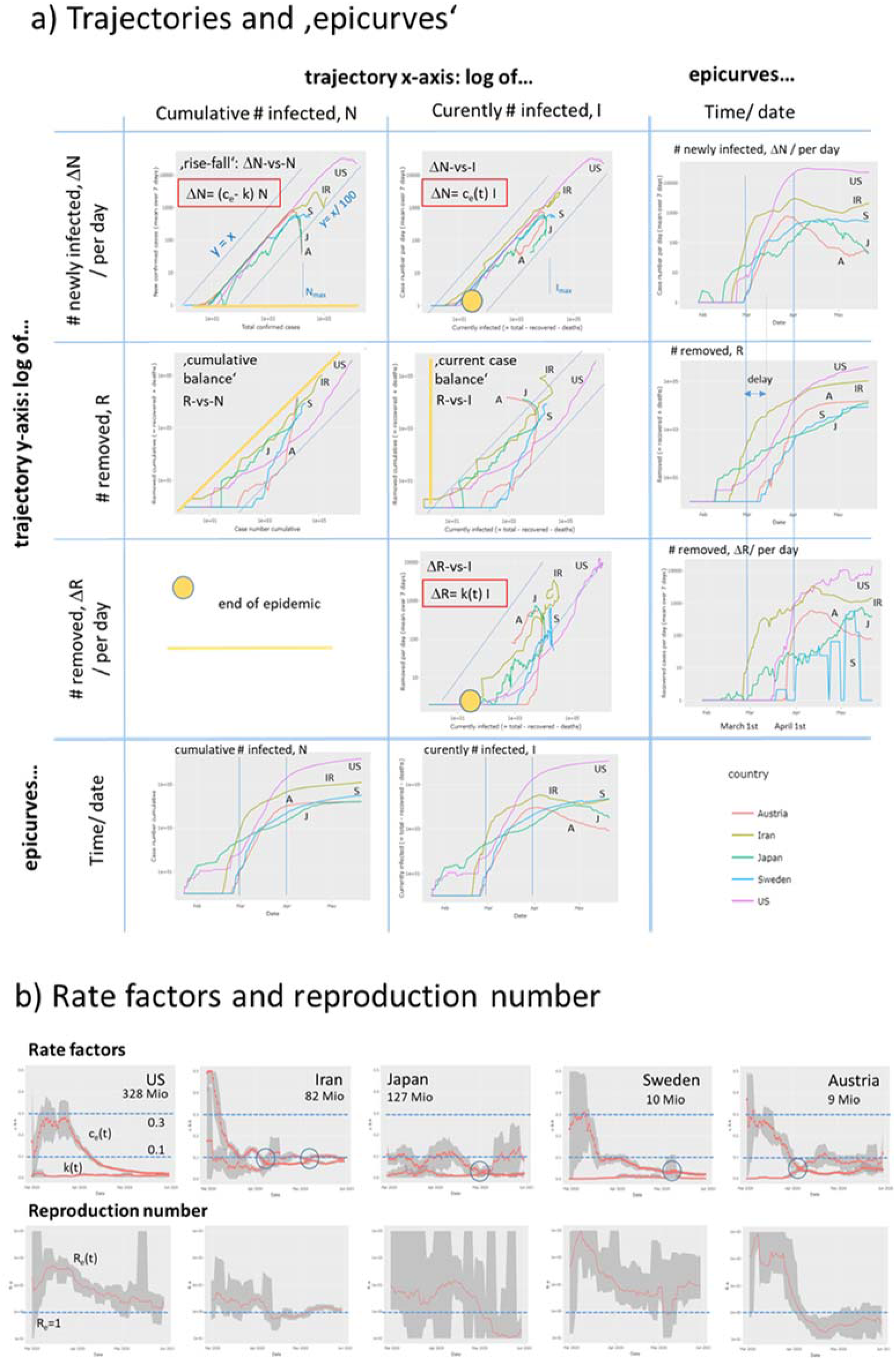
Dynamic Covid-19 characteristics of selected countries. a) Trajectories and the respective time dependencies (‘epicurves’) plot different numbers as indicated in the figure. The yellow marks (circle and lines) indicate the ‘end’ points where the trajectories are expected to return upon eradication. See also Appendix II for details. b) Rate factors of transmission and the respective reproduction numbers as a function of data.

### Monitoring Covid‐19 by reproduction numbers

Effective reproduction numbers as shown in Figure 5b provide suited summary measures of the case numbers with a well-defined epidemiological meaning. Their value defines the transmission potential in the population in terms of the mean number of individuals who are infected by one infectious person on the average. For the comparison of all or a selection of the countries available, the monitoring tool generates a ranked boxplot of their actual reproduction numbers. Presently, the epidemic is not stopped in roughly 50% of all countries because their R_e_ is still larger than unity (Figure 6). The tool also generates the respective plot for R_e_-values obtained two and four weeks before. At the latter date about 70% of countries show R_e_> 1, which demonstrates the presently decaying trend. Time courses of a selection of countries illustrate different types of decays which eventually relate to the type of NPIs taken. For example, early, consequent eradication of epidemic in Island and Croatia result in fast and steep decays. Slower but monotonous decays were observed in Russia, Spain and Portugal. Also wave-like changes before the final decay (Japan, Singapore) or even worsening of situation (Armenia, Sweden until middle of May) were found. Presently (May, 25^th^) Sweden shows the highest reproduction numbers among all countries studied. Note, that R_e_ is a relative measure considering daily changes and current numbers of infections and recoveries (Appendix I), meaning that restricted outbreak clusters affecting only relatively small numbers of individuals suggest spread of epidemic in a larger, not affected population. Hence, a combination of charcteristic numbers should be used to characterize dynamic of epidemic, namely transmission rate factor (or doubling time of cases) more at the beginning, effective reproduction numbers more in the phase of vast spread of epidemic and absolute numbers of new cases in the phase of mitigation and near eradication.

**Figure 6:**
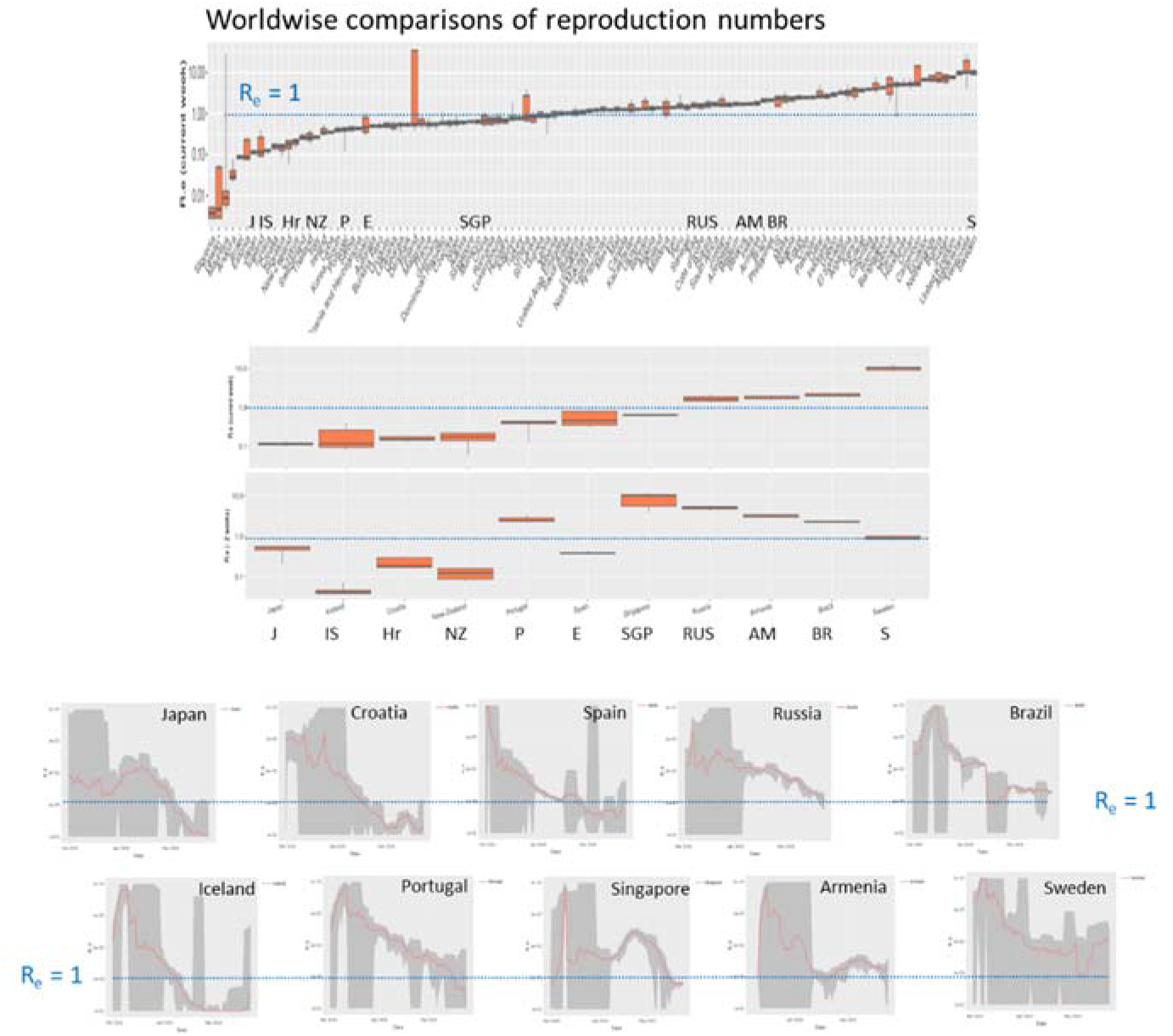
Effective reproduction numbers across the world. Ranked plot of current R_e_ values across all countries studied indicate that less than half of them fall below the epidemic threshold of R_e_=1 (horizontal dashed line). Time series for selected countries are shown as examples in the part below. They reveal sharp decays, virtually invariant and even increasing courses.

### Monitoring the effect of NPIs across Europe

Next, we asked how the NPIs taken in Middle and Western Europe and Scandinavia in the first three weeks of March 2020 affected the dynamics of the epidemic. The rise-fall trajectories of countries selected from [9] reveal that they now are mostly in the falling regime however with modifications such as parallel downwards shifts, wave-like decays and even lacking decays as already discussed above (Figure 7**Fehler! Verweisquelle konnte nicht gefunden werden.a). In Figure 7 Fehler! Verweisquelle konnte nicht gefunden werden**.b we re-plot the trajectories separately for each country together with marks indicating which measure was taken when along the trajectories. In most cases, trajectories start turning downwards about two weeks after a complete lockdown in the respective country. Before this, one often finds slowing down of the exponential growth as indicated by small differences compared with the trajectory of US referring to exponential growth. In Norway, Denmark, and also Sweden one observes a relatively strong first slowing down as indicated by the parallel downwards shift of the trajectories which roughly refers to a reduction of the transmission rate constant by about 30 % (Figure 7d). Sweden, without complete lockdown measures, but also Great Britain show weakest decay of the trajectories and largest values of the effective reproduction numbers R_e_> 1 in contrast to all other countries except Belgium (Figure 7c). Comparison of the reproduction numbers two and four weeks earlier indicates consistent high values in Britain and Sweden and also a delayed decay in Italy, Spain and France, the European countries, which were heavily hit by Covid-19 in February and March. The time courses of the reproduction numbers R_e_(t) respond nearly immediately on the measures in many cases showing, at least, small drops in support of a recent study [9] which assumes that the reproductive number – a measure of transmission – immediately responds to interventions being implemented (Figure 7d, the first measure and complete lockdown were indicated). Consistent decays to values R_e_< 1 after about two weeks were seen in Scandinavia (except Sweden) and Austria, Switzerland and Germany while in Belgium, France, Italy and Spain the decays last roughly four weeks until they fall below the epidemic threshold. In Sweden and Great Britain, virtually unchanged levels of R_e_ above the ET were observed. A recent model analysis of the effect of NPIs in Germany applies a SIR model with changed rate constants at so-called ‘change points’ which are assumed to take place when measures were applied [23]. We found a decay of the transmission rate during the time when measures were applied which drops overall by 60 – 70% in rough agreement with [23] (Figure 7d, right part). Interestingly, Japan showed a similar resonse to NPIs as the European countries, namely a slow, but instantaneous growth of epidemic turned into the falling regime at the beginningf May (Figure 5), two-three weeks after NPI measures were intensified at April 10^th^.

**Figure 7:**
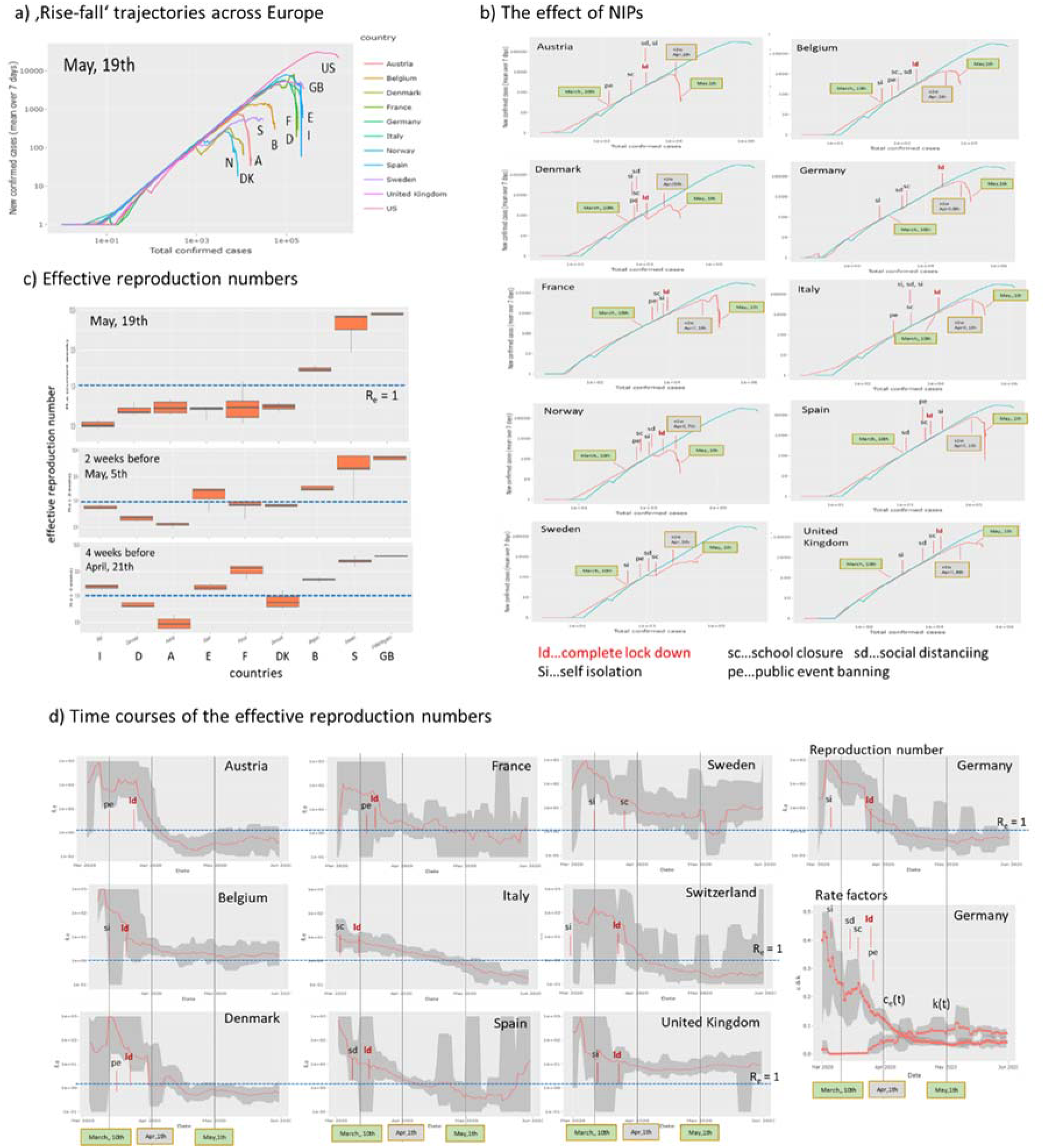
The effect of Non-Pharmaceutical Interventions across ten European countries selected in analogy with [9]. The trajectory of US is shown for comparison. a) Rise-fall trajectories of selected countries mostly decay thus indicating marked decrease of epidemic in most cases. b.) Country-by-country plots of the rise-fall trajectories together with marks assigning the NIPs (dates and assignments were taken from [9]) show that the trajectories turn downwards about two weeks after lockdown in most cases (the grey box refers to the ‘two weeks after the last measure’ date). Exceptions are Sweden (no complete lockdown) and United Kingdom. The two green boxes indicate the data obtained at March 10^th^ (mostly before measures) and May 1th. c) The effective reproduction numbers are still clearly above the critical value of R_e_=1 for Sweden and Great Britain. Italy and France show the strongest decay of R_e_ over the last 4 weeks. d) Courses of the effective reproduction number as a function of time indicate a marked drop of Re(t) immediately after the lock down in most countries. Also the first measure taken is indicated. For Germany all 5 measures were indicated together with the courses of the rate factors.

Overall, our simple analysis reveals that NPIs were followed by drops of the reproduction number mainly due to a decay of the transmission rate factor and by halt of epidemic after two to four weeks after complete lock down. Sweden (and partly GB) shows also a drop of R_e_ and the transmission rate which however overall are insufficient until end of May to stop epidemic. Both, the time-course of reproduction number and the rise-fall trajectory are sensitive to detect the slowing down and the halt of epidemic. The available country-wise numbers used do not allow to analyse the observed effect assuming heterogeneous effects of NPI on different subpopulations which, in principle, could explain steps and wavelike changes in the courses of the trajectories as an alternative to alterations of the rate factors in a homogeneous population assumed here.

### Monitoring Covid‐19 across Germany

Countries across the world differ in many factors related to Covid-19 epidemic such as the particular NPI measures, social behaviour, family structure and education systems with differing school rules, population densities, urban structure, transport system and also age distribution. The heterogeneity of these factors is assumed to be smaller inside each of the countries than between them. For Germany, the Covid-19 viewer provides ‘rise-fall’ trajectories for all sixteen German states, which cover population sizes between about 0.7 Mio (Bremen) up to 18 Mio inhabitants (Nordrhein-Westfalen). They include three city-states (Hamburg, Bremen, Berlin), while the other states are ‘area’-states include countryside regions and towns of different sizes. The trajectories overall express very similar courses of the epidemic across Germany (Figure 8a), which suggest relatively similar dynamics of the epidemic in different parts of the country and, particularly, that Germany-wide NPIs ‘locked-down’ epidemics in the different states in a similar way. Analysis of the maximum cumulative number of infected individuals, N_max_, using fits of the ‘rise-fall’ trajectories however reveals considerable differences especially between the South and West of Germany and its East and North (Figure 8b). In Bavaria, which is located in the South of Germany, roughly eight-times more people are infected on relative scale than in Mecklenburg-Vorpommern located in the North-East. In general, ‘area’ states from the West and South of Germany were more affected by epidemic than states in the East and North. This difference associates with an earlier outbreak in the former states with higher amounts of infected individuals (Figure 8c). NPIs were taken Germany-wide at the same time between 9^th^ and 23^th^ of March, which suggests that delayed measures with respect to the outbreak will increase the burden of infections. In summary, Germany-wide the trajectories reflect similar dynamics of epidemic where however earlier outbreaks especially in the West and South and in larger cities gives rise to increased numbers of infected persons, possibly because of the delay of NPI.

**Figure 8:**
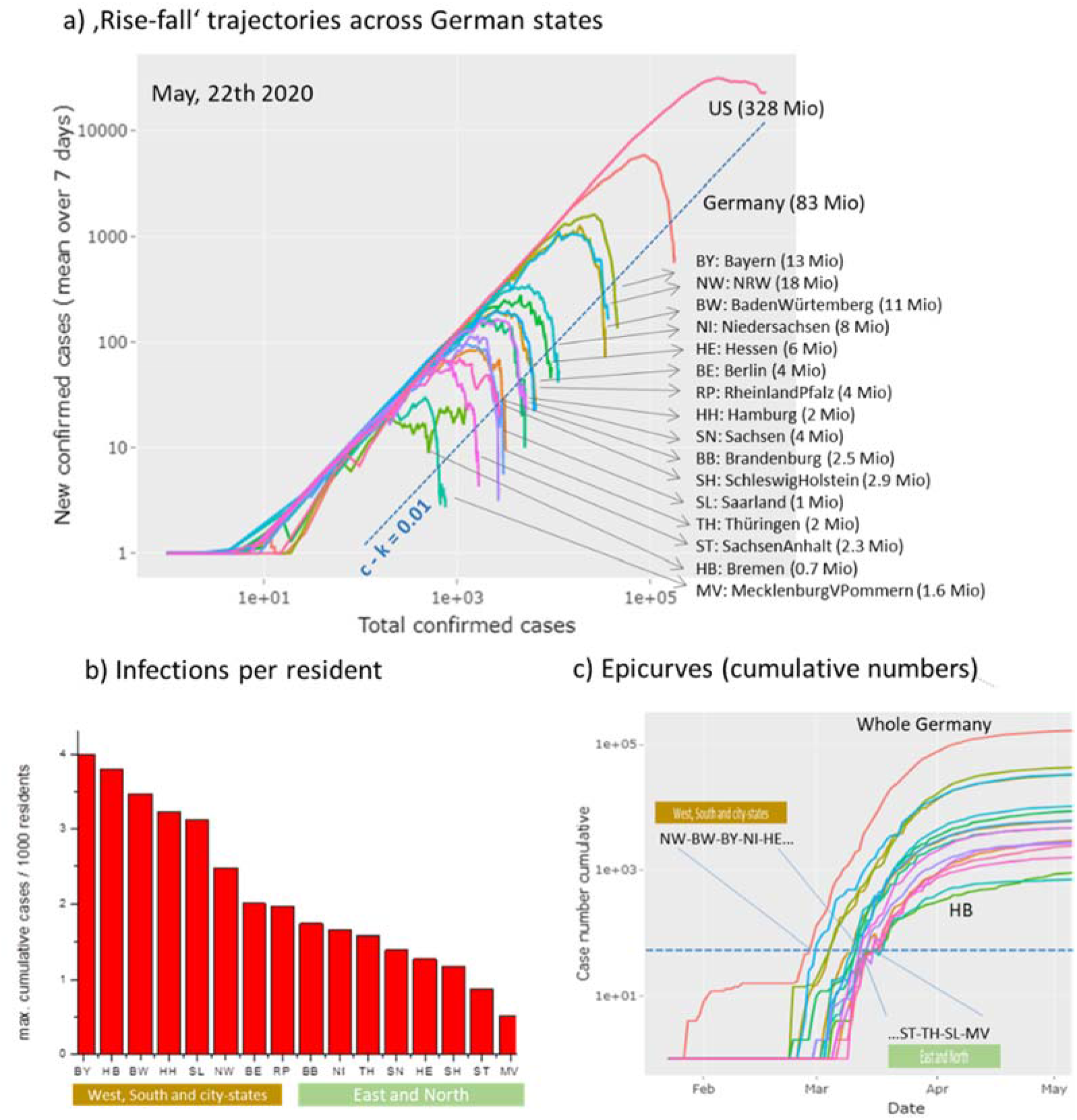
Covid-19 in Germany: a) ‘Rise-fall’ trajectories across German states: The trajectories of German states resemble that of whole Germany indicating similar dynamics of Covid-19 across Germany. Trajectory of US is shown for comparison. b) The relative maximum cumulative number of infected cases (per 1,000 residents of the respective states divides clearly into states from West and South Germany and states in the East and North of Germany. City states (BE, HH, HB) are found in the former group. c) Epi-curves (cumulative case numbers as a function of time) reveal that epidemic arrived earlier in Western and Southern states mostly by a few days compared with the Eastern and Northern ones. The curve of the city state Bremen (HB) slightly differs from that of the other ones.

### Monitoring death tolls

Mortality is an important endpoint of Covid-19 epidemic related to a series of factors such as the intrinsic severity of the virus [24] in first instance, but also age, sex, genetic and immunological predisposition [25], disease history and also co-morbidities of the patients [26], as well as the effectivity of medical measures such as ICU services [27], the capacity of health care systems and also socio-economic factors. Censing of deaths, e.g. by counting Covid-19 positively tested deaths as Covid-19 caused or not, is another factor affecting the reported numbers. So far we subsumed the numbers of death cases together with recovered individuals as removed ones. Separate counting shows that, overall, the dead toll of Covid-19 ranges from less than 1% up to more than 10% of counted infections, depending on country and time, when the data were registered (see below). The ‘custom trajectory’ page provides ‘mortality trajectories’ in terms of cumulative death cases versus cumulative infections (alternatively one can choose daily cases). Constant percentages refer to parallel diagonal lines as indicated (‘iso-percentage’ lines). For illustration we selected groups of countries in Figure 9 for comparison with part of the ‘rise-fall’ trajectories in Figure 4b. Larger-size West-European countries (Great Britain, France, Spain, Italy) all show similar mortality trajectories referring to about 10% of the (visible) infected individuals. Mortality of Germany and Austria is smaller (about 4%), possibly due to the smaller mean age of infected persons at the beginning of epidemic. The respective mortality-trajectories however slowly grow in direction of the level of the other European countries with increasing number of infections. Note that the slopes of the trajectories of the latter countries (e.g. France, Italy, Spain) is slightly steeper than that of the iso-percentage lines which indicates slowly growing mortalities across in these countries. Presently, mortality in Europe is largest in Sweden, Belgium, Netherlands and Great Britain with further increasing trends. Relative small mortality is found in Russia and Belarus possibly caused by governmental-control about Covid-19 related death-census. Mortalities in Ukraine and Estonia and in East Asia (China, Japan) are comparable with mortality in US, where the latter Asian countries and also South Korea show an increasing trend of mortality. In South America, one finds higher mortalities than in US with further increasing trends. Overall, comparison of the mortality trajectories reveals systematic differences and trends, which need further analysis for interpretation.

**Figure 9:**
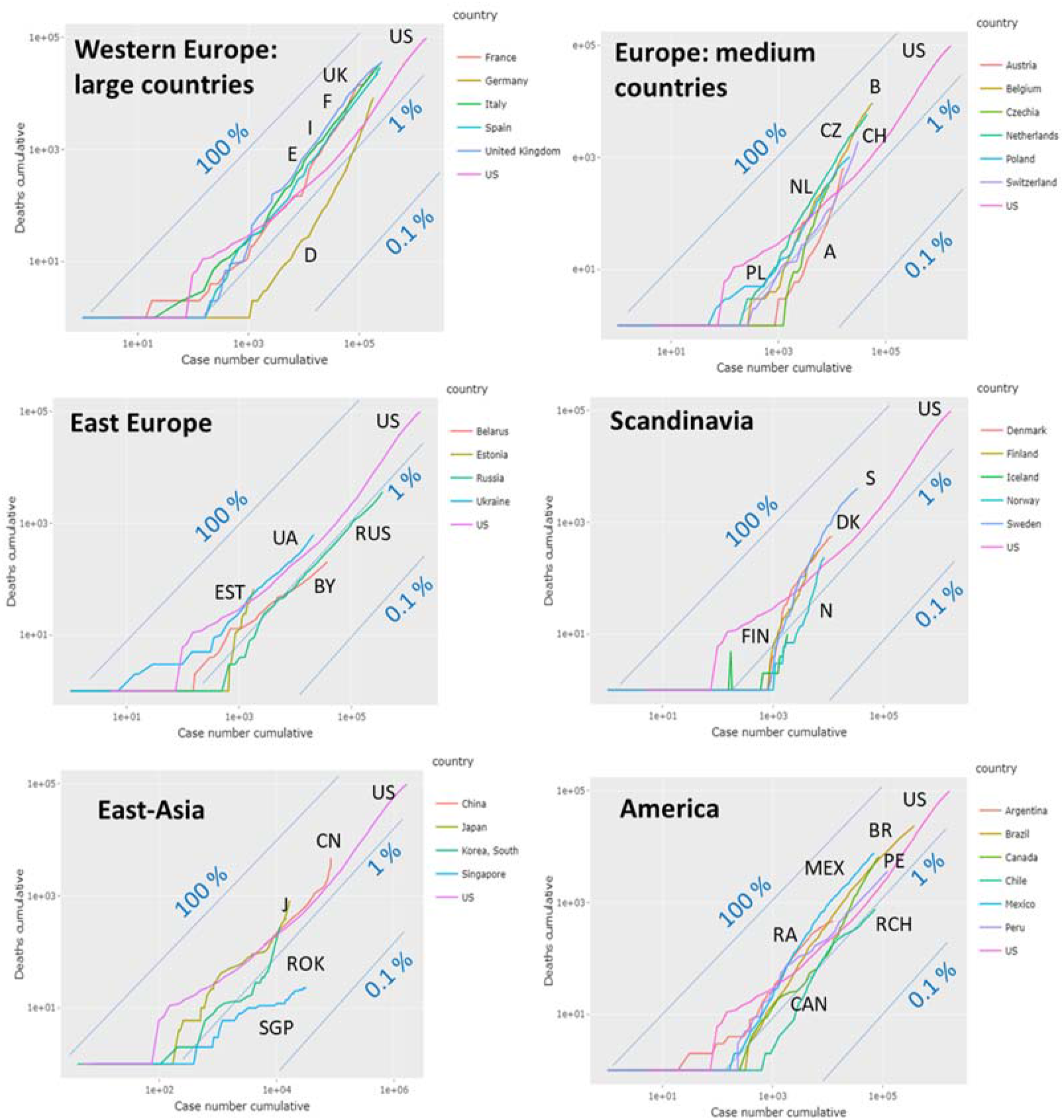
Mortality trajectories show cumulative number of deaths versus cumulative number of infections. The diagonal line (y = x) refers to 100%, parallel lines to 1% and 0.1% (number of deaths per 100 cases). The mortality trajectory of US is shown in all plots for comparison.

## Discussion

We here presented the ‘Covid-19 trajectory viewer’, which generates a series of trajectories and plots based on public available Covid19 data. It enables the comparison between epidemic development with country-wise resolution worldwide. Trajectories are based on two types of counts, namely the number of infected and of removed (recovered and died) individuals. Plots use either these counts directly, their cumulative values or increments per day and combine them in different ways, which allows to inspect the actual state of the epidemic from different perspectives.

In addition, the monitoring tool enables calculation and visualization of derived parameters, namely the effective transmission and recovery rate factors and the effective reproduction number. They estimate the transmission and removal ‘power’ as basic characteristics showing whether epidemic growths or declines. Changes of these parameters during epidemic development reflect different factors affecting the dynamic of epidemic, namely (i) the possible consequences NPIs, (ii) eventually growing immunity due to decaying numbers of susceptible individuals, and, (iii) also differences in the methods of counting and reporting data between different countries. Our monitoring metric is sensitive for subtle alterations of the dynamics of the epidemic making it suitable to estimate the effectivity of NPIs and to serve as ‘seismometer’ for secondary outbreaks to early indicate such events (Figure 10).

**Figure 10:**
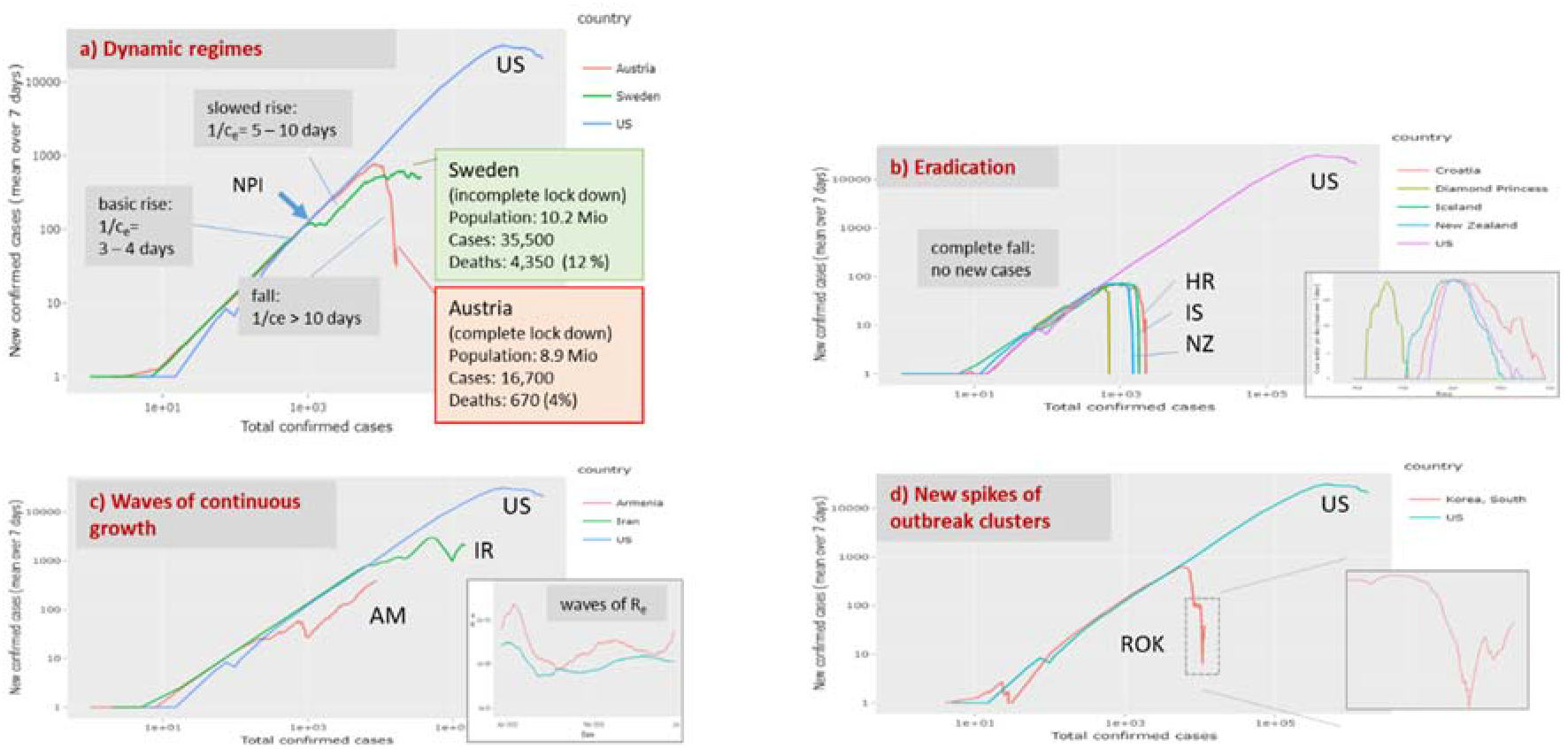
Example trajectories indicating different dynamic regimes of Covid-19: a) Basic rise and falling regimes refer to transmission intervals of 3-4 and more than 10 days, respectively. They were observed in European countries under complete lock down such as Austria. Incomplete lock down as applied in Sweden only slowed down spread of epidemic. It associates with roughly two times more infections and a more than four-fold death-toll. b) Eradication of epidemic can be expected in island states (Iceland, New Zealand) and other relatively small countries (e.g. Croatia) showing disappearance of new cases two to three months after the outbreak (see epi-curves in the insertion). Another example of eradication is Covid-19 spread at the Princess Diamond cruise liner with about 760 infections. c) Wave-like up and downs of epidemic were observed in Armenia and Iran. The trajectories transform into wave-like oscillations of the effective reproduction number (insertion). d) A new spike of cases is seen in the trajectory of South Korea. The trajectory for US is shown for comparison.

Three possible future pandemic scenarios for Covid-19 dynamics have been suggested based on previos influenca courses [1], firstly, ‘peaks and valleys’ where the first big wave in spring 2020 is followed by repetetive smaller waves with geographic specifics depending on local NPIs; secondly, the ‘fall peak’ suggesting a large secondary peak in fall, winter 2020; and, third, a ‘slow burn’ of ongoing transmission and case occurrence, but without a clear wave pattern, again with geographic variations affected by the degree of mitigation measures in place in various areas. One or none of them, or even all three in parallel in different countries will be possible, where trajectories and rate factor curves will provide an instrument to distinguish the different scenarios.

Thereby, one has to keep in mind that these are data on visible, symptomatic Covid19 cases. Un-symptomatic cases remain usually undetected and can exceed the number of symptomatic ones considerably. A recent publication shows that more than 80% of all positively tested Covid-19 cases on a cruise liner did not show any symptoms, raising questions about the true prevalence of “silent” infections [28] with possible consequences for the immunization dynamics in a population. Moreover, our simple monitoring does not explicitly consider heterogeneities of the spread of the epidemic in a population (e.g. cities versus countryside, elderly versus younger, hospitalized versus non-hospitalized, symptomatic versus asymptomatic, highly exposed professions versus less exposed ones, etc.). Such effects are hidden in the data and can be considered in terms of the trajectory approach by using more detailed data, e.g. by stratifying populations geographically, with respect to professions, age, symptoms etc. and/or by applying more elaborated models.

Our visualization in terms of trajectories and derived rate factors and their interpretation is based on the simple SIR model dividing the visible population into three types of individuals. Such three-state models have been widely and successfully used in many areas of sciences to describe different kinds of dynamics, ranging from elementary reaction kinetics in chemistry to photo-physics, molecular transformations in biology and many other fields. The basic assumption behind the SIR model is the mass action law, claiming that changes of the population of a state directly relates to its population number. The different trajectories visualize this relationship by plotting changes of newly infected or removed individuals as functions of the number of cumulative or currently infected individuals. The double logarithmic scaling of the axes accounts for the fact that the solution of the ordinary differential equations behind the SIR model predicts exponential dynamics in important limiting regimes such as the early or late outbreak limits, which in turn, suggest linear courses of the trajectories. This way the obtained trajectories reflect a virtually common maximum transmission rate in the exponential growth phase in many countries suggesting that each infected individual infects another one every two-three days (Figure 10a). The initial growth is followed by down-steps and parallel shifted lines indicative for exponential growth with reduced transmission rate (e.g., transfer of infection between two individuals every five days). Downturns of different sharpness indicate markedly reduced spread dynamics, and also halt of the epidemic in terms of falling courses if transmission frequency reaches a level of more than one per ten days.

The close temporal relatedness between slowing down of the transmission dynamics and the dates when measures of the NPI-type were taken suggests causal relations and shows that an associated ‘falling’ regime can be monitored using the trajectory approach. The NPI result in dropping transmission rates and reproduction numbers where the steepness of decay in Europe is larger for countries such as Austria and Germany, which were hit by the main infection wave a few weeks later than Italy, France and Spain showing slower decays. Early NIPs on a relative time scale with respect to growth dynamics obviously facilitate faster slowing down afterwards. So-called ‘complete lockdown’ measures seems to be an essential measure for stopping epidemic despite considerable differences between countries, e.g. in handling go-out restrictions (‘Ausgangssperre’, relatively moderate rules in Germany versus strong ones in Italy, Spain and France). The Swedish model seems to fail regarding transmission dynamics conceding further expanding epidemic and high death toll (Figure 10a). Our trajectories show that lowering a of transmission rates by more than 50-70% compared with its maximum, intrinsic value, is required to stop epidemic and to turn it into the decaying regime.

Joint plotting of trajectories using the Covid-19 viewer shows that at present majority of East Asian (China, South Korea, Singapore) and European countries are in the falling regime, while most American countries are in the exponential growth phase. Epidemic seems virtually eradicated in the island states New Zealand, Iceland but also other small countries such as Croatia in a similar way as observed at ‘Diamond Princess’ Cruise Liner held under isolation (Figure 10b). It shows that isolation in combination with strong NPIs effectively stop epidemic. On the contrary, slowing down but still exponential growth are seen in other small and relatively isolated countries such as Armenia (surrounded by mountains and closed borders to part of neighbouring countries) reflecting inefficiency of measures taken. Wavelike up and down as seen for Iran indicate repeated waves of growing epidemic (Figure 10c). New outbreak clusters become evident as spiked upturns in the falling regime as indicated presently for South Korea (Figure 10d).

The removal rate factor is a second, important characteristic of Covid-19 dynamics, which additively composes of recovery and death rates, where the former number is dominating. Removal rates can differ by a factor of two-to-ten between different countries (e.g. Germany and Austria versus Spain and France, Figure S 3) by unknown reasons. Possible explanations are specifics of the recovery process due to healthcare measures applied and/or epidemiological factors such as age- and/or health-risk of the respective populations. Also counting criteria of recovered individuals are another, possibly more relevant factor, which can differ between countries. Often census agencies apply recovery counting algorithms (e.g. by assuming recovery two weeks after infection if no other information is available in Germany) presumably biasing estimation of removal rate factors. Moreover, also counting of deaths is census-dependent. On the other hand, the initially low but afterwards increasing mortality rates in Austria and Germany can be rationalized by the increasing age of infected individuals (disease was initially spread in communities of younger persons). Thus, comparing trajectories supports detection of differences of recovery and mortality rates between countries for subsequent analysis of the possible reasons.

## Conclusions

Covid-19 pandemic develops in different phases around the world ranging from exponential growth to decaying regimes and even eradication from region to region and from country to country. It is characterized by high dynamics, which necessitate prompt monitoring to evaluate the outcome of NPI measures in either, ‘lockdown’ or ‘lock up’ direction to indicate improvement or worsening in terms of suited metrics such as increasing or decreasing numbers of cases, rate factors or reproduction numbers. The Covid-19 viewer provides this information in the worldwide context on a daily actualized basis. We understand our report as a worked example reflecting aspects of the pandemic in May 2020, which supports future monitoring using the Covid-19 viewer as a sort of working instruction. Many aspects of the Covid-19 pandemic are not completely understood. This includes dark figures of infections, detailed spreading mechanisms and associated socio-economic, politic and health factors. Here more studies reasoning differences between regions and countries are required. The trajectory approach complements epi-curve reporting by bridging the gap to modelling methods. Inspection and comparison of the trajectories and of the time courses of rate factors extracted are expected to inspire development of substantiated hypotheses and elaboration of improved models to better understand mechanisms of epidemic spread and decay and theirs specific in different countries and regions.

## Data Availability

Data were provided by https://systems.jhu.edu/research/public-health/ncov/ and https://www.rki.de/DE/Content/InfAZ/N/Neuartiges_Coronavirus/Fallzahlen.html
The tool is available via the websites of IZBI (www.izbi.de) and the Leipzig Health Atlas (https://www.health-atlas.de/models/28).

## Subject area

Health Informatics

## Author approval

All authors have seen and approved the manuscript.

## References

1. Kristine A. Moore, Lipsitch M, John M. Barry, Osterholm MT: COVID-19: The CIDRAP Viewpoint: Part 1: The Future of the COVID-19 Pandemic: Lessons Learned from Pandemic Influenza. https://www.cidrapumnedu/sites/default/files/public/downloads/cidrap-covid19-viewpoint-part1_0pdf 2020.

2. Hopkins J: Coronavirus Ressource Centre. https://coronavirusjhuedu/ 2020.

3. Roser M, Ritchie H, Ortiz-Ospina E, Hasell J: Coronavirus Pandemic (COVID-19). In: *Published online at OurWorldlnDataorg.* vol. https://ourworldindata.org/coronavirus; 2020.

4. Times F: Coronavirus tracked: the latest figures as countries fight to contain the pandemic. Financial Times 2020, https://www.ft.com/content/a26fbf7e-48f8-11ea-aeb3-955839e06441.

5. The COVID tracking project. https://covidtrackingcom/ 2020.

6. Bock W, Adamik B, Bawiec M, Bezborodov V, Bodych M, Burgard JP, Goetz T, Krueger T, Migalska A, Pabjan B et al: Mitigation and herd immunity strategy for COVID-19 is likely to fail. *medRxiv* 2020:2020.2003.2025.20043109.

7. Barbarossa MV, Fuhrmann J, Heidecke J, Vinod Varma H, Castelletti N, Meinke JH, Krieg S, Lippert T: A first study on the impact of current and future control measures on the spread of COVID-19 in Germany. *medRxiv* 2020:2020.2004.2008.20056630.

8. Donsimoni JR, Glawion R, Plachter B, Waelde K: Projecting the Spread of COVID19 for Germany. *medRxiv* 2020:2020.2003.2026.20044214.

9. Seth Flaxman SM, Axel Gandy, H Juliette T Unwin, Helen Coupland, Thomas A Mellan, Harrison Zhu, Tresnia Berah, Jeffrey W Eaton, Pablo N P Guzman, Nora Schmit, Lucia Callizo, Imperial College COVID-19 Response Team, Charles Whittaker, Peter Winskill, Xiaoyue Xi, Azra Ghani, Christl A. Donnelly, Steven Riley, Lucy C Okell, Michaela A C Vollmer, Neil M. Ferguson, Samir Bhatt: Estimating the number of infections and the impact of non-pharmaceutical interventions on COVID-19 in European countries: technical description update. *arXiv:200411342* 2020.

10. Khailaie S, Mitra T, Bandyopadhyay A, Schips M, Mascheroni P, Vanella P, Lange B, Binder S, Meyer-Hermann M: Estimate of the development of the epidemic reproduction number Rt from Coronavirus SARS-CoV-2 case data and implications for political measures based on prognostics. *medRxiv* 2020:2020.2004.2004.20053637.

11. Dandekar R, Barbastathis G: Quantifying the effect of quarantine control in Covid-19 infectious spread using machine learning. *medRxiv* 2020:2020.2004.2003.20052084.

12. Biswas K, Khaleque A, Sen P: Covid-19 spread: Reproduction of data and prediction using a SIR model on Euclidean network. *arXiv:200307063* 2020.

13. Engbert R, Rabe MM, Kliegl R, Reich S: Sequential data assimilation of the stochastic SEIR epidemic model for regional COVID-19 dynamics. *medRxiv* 2020:2020.2004.2013.20063768.

14. Maier BF, Brockmann D: Effective containment explains subexponential growth in recent confirmed COVID-19 cases in China. Science 2020:eabb4557.

15. Soucy J-PR, Sturrock SL, Berry I, Westwood DJ, Daneman N, MacFadden DR, Brown KA: Estimating effects of physical distancing on the COVID-19 pandemic using an urban mobility index. *medRxiv* 2020:2020.2004.2005.20054288.

16. Chen Y-CL, Ping-En; Chang, Cheng-Shang; Liu, Tzu-Hsuan: A Time-dependent SIR model for COVID-19 with Undetectable Infected Persons. *eprint arXiv:200300122* 2020.

17. Jonas Dehning JZ, F. Paul Spitzner, Michael Wibral, Joao Pinheiro Neto, Michael Wilczek, Viola Priesemann: Inferring change points in the COVID-19 spreading reveals the effectiveness of interventions. *arXiv:200401105* 2020.

18. Chernyshev A: Autocatalytic Model for Covid-19 Progression in a Country. *medRxiv* 2020:2020.2004.2003.20052985.

19. Pell B, Kuang Y, Viboud C, Chowell G: Using phenomenological models for forecasting the 2015 Ebola challenge. Epidemics 2018, 22:62-70.

20. Wangping J, Ke H, Yang S, Wenzhe C, Shengshu W, Shanshan Y, Jianwei W, Fuyin K, Penggang T, Jing L et al: Extended SIR Prediction of the Epidemics Trend of COVID-19 in Italy and Compared With Hunan, China. Frontiers in Medicine 2020, 7(169).

21. Su L, Hong N, Zhou X, He J, Ma Y, Jiang H, Han L, Chang F, Shan G, Zhu W et al: Evaluation of the Secondary Transmission Pattern and Epidemic Prediction of COVID-19 in the Four Metropolitan Areas of China. Frontiers in Medicine 2020, 7(171).

22. W Chang, J Cheng, J Allaire, Y Xie, McPherson J: Shiny: web application framework for R. R package version 2017.

23. Dehning J, Zierenberg J, Spitzner FP, Wibral M, Neto JP, Wilczek M, Priesemann V: Inferring change points in the spread of COVID-19 reveals the effectiveness of interventions. Science 2020:eabb9789.

24. Yinon M Bar-On AF, Rob Phillips, Ron Milo: SARS-CoV-2 (COVID-19) by the numbers. eLife 2020, eLife 2020;9:e57309.

25. Casanova J-L, Su HC: A global effort to define the human genetics of protective immunity to SARS-CoV-2 infection. Cell 2020.

26. Petrilli CM, Jones SA, Yang J, Rajagopalan H, O’Donnell LF, Chernyak Y, Tobin K, Cerfolio RJ, Francois F, Horwitz LI: Factors associated with hospitalization and critical illness among 4,103 patients with COVID-19 disease in New York City. *medRxiv* 2020:2020.2004.2008.20057794.

27. Docherty AB, Harrison EM, Green CA, Hardwick HE, Pius R, Norman L, Holden KA, Read JM, Dondelinger F, Carson G et al: Features of 16,749 hospitalised UK patients with COVID-19 using the ISARIC WHO Clinical Characterisation Protocol. *medRxiv* 2020:2020.2004.2023.20076042.

28. Ing AJ, Cocks C, Green JP: COVID-19: in the footsteps of Ernest Shackleton. Thorax 2020:thoraxjnl-2020-215091.

29. Ma J: Estimating epidemic exponential growth rate and basic reproduction number. Infectious Disease Modelling 2020, 5:129-141.

30. Britton T, Trapman P, Ball FG: The disease-induced herd immunity level for Covid-19 is substantially lower than the classical herd immunity level. *medRxiv* 2020:2020.2005.2006.20093336.

31. Fraser C: Estimating Individual and Household Reproduction Numbers in an Emerging Epidemic. PLOS ONE 2007, 2(8):e758.

32. Chen Y-C, Lu P-E, Chang C-S, Liu T-H: Time-dependent SIR model for COVID-19 with Undetectable Infected Persons. *arXivorg* 2020, arXiv:2003.00122.

